# Research and learning priorities for a surgical obstetrics and family planning project implementing in low- and middle-income countries: results of an expert consultation

**DOI:** 10.1101/2025.05.03.25326922

**Authors:** Farhad A. Khan, Karen Levin, Renae Stafford, Vandana Tripathi

**Author notes:** **Role of authors** FAK, KL, RS, and VT designed and implemented the consultation. FAK, KL, RS, and VT developed the data collection instrument. FAK and VT planned data analysis. FAK oversaw data collection and analysis. FAK, KL, RS, and VT prepared the first draft of this manuscript. All authors reviewed and edited drafts of this manuscript and approved this version for submission.

## Abstract

**Introduction:** Cesarean delivery, peripartum hysterectomy, female genital fistula treatment, and long acting and permanent contraceptive method provision comprise an important set of surgical procedures in reproductive and maternal health. The volume of these procedures is growing in low- and middle-income countries (LMIC). Establishing research priorities in a learning agenda for surgical obstetrics and family planning represents a key step in generating and using evidence to improve health outcomes associated with these surgeries.

**Methods:** A safe surgery project addressing family planning and obstetrics used a two-stage rating and ranking consultation process to prioritize topics in its learning agenda, focusing on LMIC needs. A list of research and learning topics spanning the project’s technical areas (surgical obstetric care [cesarean delivery and peripartum hysterectomy], fistula prevention and treatment, family planning, and cross-cutting safe surgery) was curated by searching the literature, conducting project-related surveys of experts and partners, and soliciting an expert panel via virtual consultation. Topics were rated through an online survey of the experts on four criteria: feasibility, technical importance, level of saturation, and potential for impact. The expert panel then reconvened to rank and refine highly rated topics.

**Results:** A total of 39 people participated in the expert panel, representing multilateral, academic, and funding organizations, implementing partners, and professional associations active in LMIC. Fifteen topics were prioritized across the four technical areas. Prioritized topics cover themes of prevention (e.g., intrapartum/midwifery practices to prevent unnecessary cesarean), care-seeking (e.g., social behavior change strategies for fistula prevention), peri-operative care (e.g., use of quality improvement tools including checklists and audits), and post-operative care (e.g., effective measurement approaches for monitoring outcomes).

**Conclusion:** This agenda guides clinical and programmatic learning across the safe surgery ecosystem. Collaborative action across program initiatives and clinical and community settings may contribute to significant evidence building in these priority topics.

**Teaser Key Message:**

- We identified research and learning priorities on surgical obstetrics and family planning for a project’s research and learning agenda. Practitioners, program implementers, donors and researchers involved in global safe surgery, maternal health, and family planning can apply these priorities toward guiding their evidence generation and translation efforts.

**Key Findings:**

- Safe surgery is essential for reducing maternal morbidity and mortality; this consultation represents one instance in which preventative, care-seeking, perioperative, and postoperative priorities were collaboratively identified to strengthen surgical maternal health and family planning services in LMIC.

**Key Implications:**

- Although the topics prioritized in this agenda were developed in the context of a project, they provide donors interested in maternal health and family planning with lines of inquiry to target investments in research and learning.
- Practitioners, program implementers, and researchers in surgical maternal health and family planning care can fill evidence gaps by applying these lines of inquiry within their interventions and sharing learning across programs and settings.
- This agenda, in focusing on cross-cutting knowledge gaps related to safe surgery, can complement priority learning questions in national strategic plans for surgery, family planning, and/or maternal health; priorities for policy research; and broader maternal health and family planning research agendas.

## Introduction

Despite significant reductions over time, the global maternal mortality ratio remains high at 211 deaths per 100,000 live births.^1^ Sustained, accelerated reductions in maternal morbidity and mortality in low- and middle-income countries (LMICs) require universal access to quality, safe surgical services,^2^ including long acting and reversible contraceptives (LARCs) and permanent family planning methods (PMs), cesarean delivery, peripartum hysterectomy, and fistula prevention and treatment. Although significant gaps in global access to safe surgical care exist,^3^ rural facilities experience limited human and material resources whereas urban facilities experience excessive demand for services.^4^ Under both circumstances, facility readiness to perform quality, indicated surgical procedures remains an issue. These issues are particularly important in all settings as the volume of surgical procedures increases without comparable increases in capacity.^5^ These trends are concerning; an example of the consequences is the increasing iatrogenic fistula associated with cesarean delivery and hysterectomy in a number of LMICs.^6,7^

MOMENTUM Safe Surgery in Family Planning and Obstetrics is the United States Agency for International Development’s (USAID) flagship global safe surgery initiative. The project seeks to reduce maternal morbidity and mortality by supporting country health systems actors (governments, institutions, organizations) to strengthen their capacity to provide surgical services, focusing on long acting and permanent family planning methods, cesarean delivery, peripartum hysterectomy, and fistula prevention and treatment.^8^ As part of project aims to utilize evidence and adaptive learning methods, identifying knowledge gaps can serve as a key step to strengthening surgical systems, and in turn, access and quality.

A learning (or research) agenda is comprised of questions or topics aiming to address knowledge gaps pertaining to program implementation, activities designed to answer them, and resulting dissemination products.^9^ A strategically developed and implemented learning agenda can accelerate the translation of evidence into practice. For example, in a previous global project,^10^ the prioritization of iatrogenic fistula in that learning agenda was followed by incorporating etiology tracking in routine monitoring systems,^6,11^ provider surveys, a technical consultation on cesarean section safety,^12^ and ultimately a body of evidence and action contributing to focus on surgical safety. An association between gender-based violence (GBV) and fistula was observed in a study that was done in response to the need to characterize socio-cultural factors of fistula,^13^ and is currently being translated into practice by means of integrating GBV screening and referral into treatment.

Current research agendas on safe surgery in LMICs focus on areas of unmet need for surgical patients,^14^ and knowledge gaps needed for surgical service delivery and policy in Southern Africa.^15^ Disseminated research priorities in maternal, newborn, child health, and reproductive health have included fistula prevention and treatment,^11^ sexual reproductive health and rights for the World Health Organization (WHO) African Region,^16^ family planning in Uganda,^17^ Niger,^18^ and Mozambique,^19^ and improving respectful care for newborns globally.^20^ Since obstetric, fistula, and family planning surgical services comprise important surgical interventions in LMIC,^21^ prioritizing learning topics to pursue within these technical areas is a valuable complement to other existing agendas.

We sought to develop a learning agenda, in the context of this project, to identify and prioritize topics in need of exploration across the project’s four key technical areas: surgical obstetric care, family planning (long-acting and permanent methods), fistula prevention and treatment, and cross-cutting safe surgery. This learning agenda was developed using a prioritization exercise, in which learning topics were identified and subject matter experts were convened to rate and rank learning topics.

## Methods

A variety of methods can be used to establish priorities in health research.^22^ Among these include the Delphi method, the Child Health and Nutrition Research Initiative (CHNRI) method, and consultation processes.^22^ The Delphi method involves convening a panel of experts to first solicit topics in an open-ended questionnaire followed by a series of structured questionnaires to rate and rank topics across multiple rounds. Consensus is established by presenting findings back to the expert group and asking them to adjust their ratings over multiple rounds. Common shortcomings of the Delphi method include replicability difficulties and potential for participants with strong opinions or those managing the process to influence participants ratings. The CHNRI method also involves convening an expert panel, soliciting research topics from the panel, and asking the panel to score topics according to five criteria: answerability, equity, impact on burden, deliverability, and effectiveness.^22,23^ The CHNRI method relies on the quantitative analysis of scoring data to generate a “collective” result, approximating consensus. Consultation methods involve combinations of focus group discussions and key informant interviews, though consultations reported in the literature tend to provide fewer details on their conduct.^22^

Efforts seeking to establish research priorities in family planning and maternal and newborn health have adopted and modified a variety of approaches. The CHNRI method was used by the WHO to establish a global research agenda for family planning,^24^ the WHO African Region to prioritize topics in sexual and reproductive health and rights (SRHR),^16^ and by the WHO convened expert panel to prioritize topics in maternal and perinatal health.^25^ A modified Delphi method was used by a group seeking to establish priorities in respectful newborn care.^20^ Consultation-based methods were utilized by WHO to establish priorities in SRHR for young adolescents,^26^ as well as in efforts to establish research and learning agendas for family planning in six countries.^27^ Hybrid approaches involving questionnaires and discussions attempt to take advantage of quantitative measurements from Delphi and CHNRI-based methods while also accounting for potential nuances uncovered through discussion. A hybrid approach was used to develop and validate a quality measurement index for facility-based labor.^28^ Similarly, the USAID-funded Fistula Care *Plus* project adopted a consultation-based method but utilized a questionnaire with scoring criteria (feasibility, potential for impact, saturation, and technical importance) in order establish a quantitative basis for prioritization prior to consensus-building through consultation.^11^

### Consultation Design

In order to prioritize topics for a learning agenda for surgical obstetrics and family planning programming, we solicited the opinion of an expert group through a two-stage rating and ranking consultation process, adapting and modifying the CHNRI and consultation-based approaches. Specifically, we adapted an approach from a similar research prioritization consultation deployed by the USAID-funded Fistula Care *Plus* project described above.^11^ We modified this hybrid process for the COVID context by holding two virtual consultations spaced a month apart with an online rating survey held in between. These modifications allowed us to generate research priorities rapidly, which was especially important to us because we intended to use the agenda to identify evidence generation activities to pursue in order to advance learning in surgical obstetrics and family planning over the life of the project.

We engaged the expert group in a virtual consultation meeting held in January 2022 to introduce potential learning topics and rating criteria, deployed a survey to the expert group to rate topics, and held a second virtual consultation meeting at the end of February 2022 to rank topics by technical area (figure 1).

**Figure 1.**
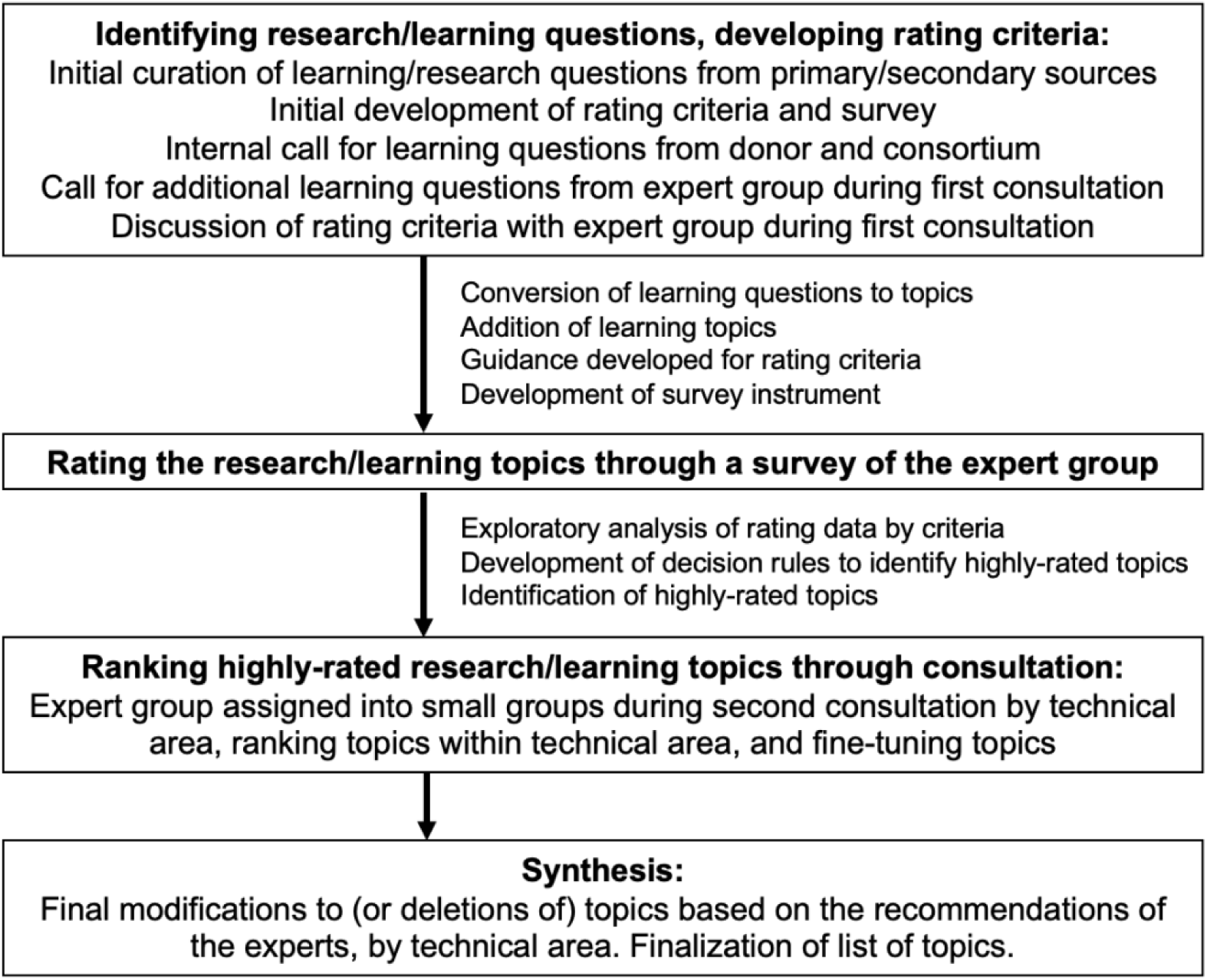
Expert Consultation Process Flow

### Expert Group

The participants engaging in this consultation were identified purposively for their expertise in at least one of the project’s four technical areas in the LMIC context: safe surgery, obstetrics, family planning, and fistula prevention and treatment. We sought out experts from academia, implementing partner organizations / international non-government organizations, the donor community, and regional professional associations (box 1). Project implementation encompasses global and country-specific activity, and consultation participants were primarily identified from organizations at the global and regional levels. As such, women and families were not engaged in this consultation. Global implementing partner organizations work with local institutions, including governments, to build demand for and capacity to provide high quality health services; while this does not include direct service provision, it often includes research. Participants were invited to participate in the consultations in December 2021, in advance of the first meeting held at the end of January 2022.

#### Box 1.

##### Institutional Affiliations of Expert Group Participants

Aga Khan University Medical College, East Africa

Bill and Melinda Gates Foundation

EngenderHealth

G4 Alliance

IntraHealth International

Jhpiego

Johns Hopkins University Center for Communications Programs

London School of Hygiene and Tropical Medicine

Population Council

Population Reference Bureau

Program in Global Surgery and Social Change, Harvard Medical School

United Nations Population Fund

United States Agency for International Development

University College London

University Gamal Abdel Nasser, Conakry, Guinea

University of Alberta

University of California, San Francisco

University of Ibadan / Center for Population and Reproductive Health

University of Minnesota

University of Zimbabwe / College of Surgeons of East, Central and Southern Africa

World Health Organization

### Identifying Research/Learning Topics and Developing Rating Criteria

A mix of primary and secondary sources were used to identify research and learning questions, including a survey of attendees of the USAID Fistula Care *Plus* End of Project Learning Event,^29^ the WHO African Region Research Priorities on Sexual and Reproductive Health and Rights,^16^ the technical application for MOMENTUM Safe Surgery in Family Planning and Obstetrics award, and a call for learning topics from core consortium partners and USAID’s Office of Maternal and Child Health and Nutrition and Office of Population and Reproductive Health. Following the initial collection of learning and research topics/questions, topics were edited to be phrased in the form of a question and all research/learning questions were categorized into one of four technical areas: cross-cutting safe surgery, surgical obstetric care, fistula repair and prevention, and family planning.

The research and learning questions were presented to the expert group during the first consultation meeting, held January 2022. Participants were asked to discuss two questions: (1) “Are there glaring gaps—do additional pressing learning questions immediately come to mind?” and (2) “Are there topics with similar questions that need to [be] combined/grouped so that a topic of great interest doesn’t end up rated/ranked lower because of a “split vote”?” Participant feedback indicated that the questions were close-ended in nature, only addressing the “what”, and neglected to understand the “how”, or “why” – i.e., pathways to impact. As a result of these discussions, the research questions that we anticipated rating were ultimately converted back into to broader learning topics, along with other additions and deletions made in response to expert group comments.

Rating criteria were adapted from a past research prioritization exercise,^11^ and included feasibility, technical importance, unsaturated topic, and potential for program impact. Ratings followed a five-point scale, with one being the lowest, and five being the highest (e.g., a rating of “1” for a topic on feasibility indicates very limited feasibility and a rating of “5” for a topic on technical importance indicates a topic of very high importance). Participants in the first consultation meeting, were asked to reflect on the criteria categories through the discussion question: “Are there other crucial criteria (other than feasibility, technical importance, lack of saturation, and potential for program impact) that you would recommend be included in the rating survey?” Based on the discussion, the criteria definitions were revised to also account for alignment with country priorities or will/commitment to pursue the topic, and ability for a topic to be applied globally (table 1).

**Table 1.**
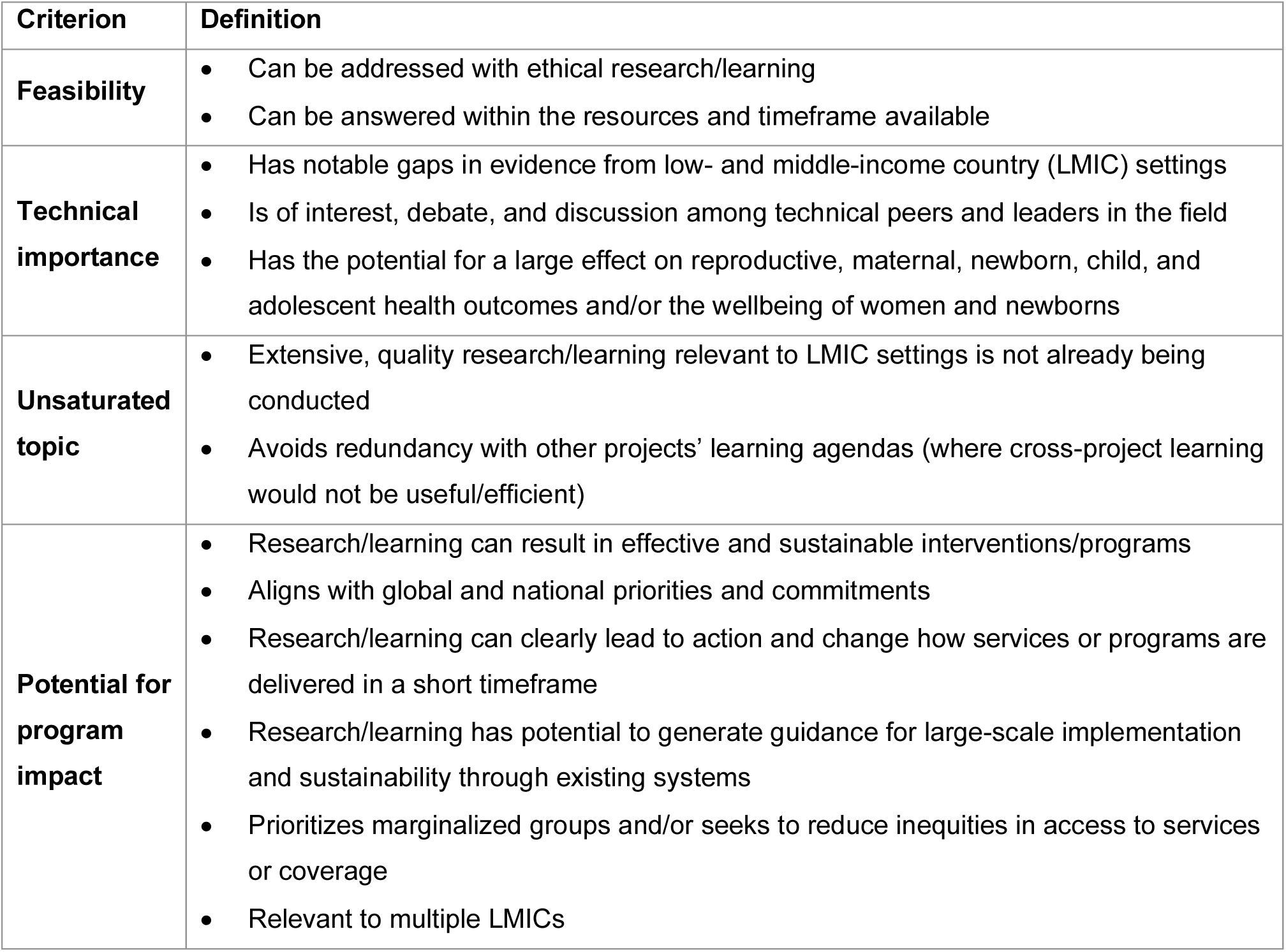
Final Rating Criteria and Definitions.

| **Criterion** | **Definition** |
| --- | --- |
| **Feasibility** | • Can be addressed with ethical research/learning<br>• Can be answered within the resources and timeframe available |
| **Technical importance** | • Has notable gaps in evidence from low- and middle-income country (LMIC) settings<br>• Is of interest, debate, and discussion among technical peers and leaders in the field<br>• Has the potential for a large effect on reproductive, maternal, newborn, child, and adolescent health outcomes and/or the wellbeing of women and newborns |
| **Unsaturated topic** | • Extensive, quality research/learning relevant to LMIC settings is not already being conducted<br>• Avoids redundancy with other projects' learning agendas (where cross-project learning would not be useful/efficient) |
| **Potential for program impact** | • Research/learning can result in effective and sustainable interventions/programs<br>• Aligns with global and national priorities and commitments<br>• Research/learning can clearly lead to action and change how services or programs are delivered in a short timeframe<br>• Research/learning has potential to generate guidance for large-scale implementation and sustainability through existing systems<br>• Prioritizes marginalized groups and/or seeks to reduce inequities in access to services or coverage<br>• Relevant to multiple LMICs |

### Rating the Research/Learning Topics through Survey

Following the first consultation meeting, the research and learning topics and rating criteria were programmed into a survey (supplementary appendix, survey instrument). The survey instrument was reviewed for usability by two members of the expert group. The purpose of this rating survey was to determine which subset of topics rated highest overall. The survey was shared with the expert group and open for responses between February 7 and 24, 2022.

Once the survey responses were collected, average ratings were generated for each criterion and each topic, across all participants. For example, an average rating for the “feasibility” of “effective strategies to strengthen key aspects of the safe surgery ecosystem” was calculated. A topic’s overall rating was the average of the average scores of each of the criterion. All criteria were weighted equally in calculation of overall ratings. Topics were then sorted from highest to lowest by overall rating. The distribution of the ratings was analyzed, and decision rules were selected to define which topics to present to the expert group as “highly rated” during the ranking stage. Outliers were analyzed to assess the extent to which the overall rating was influenced by any individual criterion. Given the overlapping nature of the technical areas, all participants were free to rate all the topics.

### Ranking Highly Rated Research/Learning Topics through Consultation

A second virtual consultation session was held at the end of February 2022 to share back which learning and research topics rated highly and for the expert group to rank these topics for prioritization in a learning agenda.

The expert group was divided into four groups, based on their self-reported expertise, among the four technical areas: cross-cutting safe surgery, surgical obstetric care, fistula repair and prevention, and family planning. Each group was shown a list of the top-rated topics meeting the decision rule and proceeded to rank the topics for priority in a learning agenda, taking all the criteria into account holistically. A facilitator managed the discussions in each of the small groups, solicited feedback from all participants, and took notes. In some circumstances, the expert group recommended rewording or merging learning topics.

The small groups also discussed possible specific research and learning questions within the topics, study designs and research methods for investigating the topics, and potential countries in which to conduct research for topics that required primary data collection. The results of these discussions are reported elsewhere.^30^

### Synthesis

Following the second consultation, refinements to topics were made to reflect the recommendations of the expert groups and the list of topics, by technical area, was finalized.

## Results

39 unique participants of the expert group made contributions across the various stages: 32 attended the first consultation; 33 responded to the survey; and 30 attended the second consultation. Of the 30 attending the second consultation, 29 participated in small groups: seven in the cross-cutting safe surgery group, eight in the fistula group, seven in the family planning group, and seven in the obstetrics group.

### Overview of Topics

A total of 63 topics were included in the rating exercise. These topics were identified following the initial curation of topics from secondary sources (e.g., peer-reviewed literature, project documentation, project-related surveys of experts and partners), the consortium of implementing partners within the project, donor, and the expert group: 18 for surgical obstetric care, 20 for fistula prevention and treatment, eight for family planning, and 17 for cross-cutting safe surgery (table 2 and supplementary tables 1-4, survey instrument in supplementary materials). The topic with the highest overall rating was “Effective strategies to strengthen key aspects of the safe surgery ecosystem (e.g., anesthesia supplies, blood and oxygen)” (supplementary table 4). On average, the cross-cutting safe surgery topics were rated highest, and the fistula prevention and treatment topics were rated the lowest, though the average and the top 25% ratings were closely distributed (table 2). Specifically, the top 25% rating for surgical obstetric care, fistula prevention and treatment, and cross-cutting safe surgery were 3.95, 3.99, and 4.07, respectively. Topics rating in the top 25% of their respective technical area were then presented to the expert group for ranking, except for family planning in which the top 5 topics were presented (tables 3-6). The top 5 family planning topics were presented for ranking because few family planning topics were curated and applying a similar “top 25%” decision rule would have yielded too few topics for discussion. The ranking exercise produced a subset of topics prioritized for inquiry (table 7).

**Table 2.** Distribution of Ratings across Surgical Obstetric Care, Fistula Prevention and Treatment, Family Planning, and Cross-Cutting Safe Surgery.

|  | **Surgical Obstetric Care** | **Fistula Prevention and Treatment** | **Family Planning** | **Cross-Cutting Safe Surgery** |
| --- | --- | --- | --- | --- |
| Number of topics | 18 | 20 | 8 | 17 |
| Minimum rating | 3.54 | 3.25 | 3.72 | 3.51 |
| Maximum rating | 4.16 | 4.15 | 4.09 | 4.35 |
| Average rating | 3.82 | 3.77 | 3.85 | 3.95 |
| Median rating | 3.78 | 3.86 | 3.82 | 3.94 |
| Third quartile (Q3) / top 25% | 3.95 | 3.99 | 3.89 | 4.07 |
| Number of topics scoring in top 25% | 5 | 6 | 2 | 5 |
| Cutoff rating / decision rule | Q3 | Q3 | Top 5 | Q3 |

### Surgical Obstetric Care

#### Rating

Of the 18 surgical obstetric topics rated, five rated in the top 25% and were presented back for ranking (table 3). Among these, the topic with the highest overall rating was “Using post-discharge/post-operative visits (in person or telehealth) to monitor post-operative morbidity and neonatal outcomes” (table 3).

**Table 3.** Surgical Obstetric Care Topics Ranked for Prioritization by the Expert Group.

| Topic | n | Rating |  |  |  |  |
| --- | --- | --- | --- | --- | --- | --- |
|  |  | Feasibility | Technical Importance | Unsaturated Topic | Potential for Impact | Overall |
| 1. Using post-discharge/post-operative visits (in person or telehealth) to monitor post-operative morbidity and neonatal outcomes | 28 | 4.25 | 4.28 | 3.86 | 4.24 | **4.16** |
| 2. Use of tools such as clinical checklists and audits for cesarean delivery (CD) and peripartum hysterectomy to enhance decision-making as part of quality improvement | 29 | 4.55 | 4.30 | 3.50 | 4.17 | **4.13** |
| 3. Intrapartum/midwifery care practices to reduce unnecessary CD | 29 | 4.07 | 4.34 | 3.55 | 4.38 | **4.09** |
| 4. Trial of labor after CD (TOLAC) or vaginal birth after CD (VBAC): LMIC practices, availability/coverage, outcomes | 27 | 4.37 | 4.30 | 3.81 | 4.19 | **4.01** |
| 5. LMIC policies and practices for subsequent pregnancies / deliveries post-CD | 28 | 3.96 | 4.00 | 4.00 | 3.93 | **3.97** |

#### Ranking and Synthesis

Consultation participants noted similarities between two topics, “Trial of labor after CD (TOLAC) or vaginal birth after CD (VBAC): LMIC practices, availability/coverage, outcomes” and “LMIC policies and practices for subsequent pregnancies/deliveries post-CD,” and elected to retain the former, which was more comprehensive.

Of the topics discussed (table 3), “Using post-discharge/post-operative visits (in person or telehealth) to monitor post-operative morbidity and neonatal outcomes” emerged as the highest priority, primarily for its potential for impact. In discussing the importance of post-discharge/post-operative follow-up, participants noted that clinicians know very little about what happens to women and newborns after obstetric surgical procedures and that such knowledge could shed light on the true level of morbidity that a client may be experiencing (e.g., surgical site infections). Participants also discussed the extent to which a follow-up study could be operationalized, highlighting that simple approaches such as phone calls may be a feasible and cost-effective way of follow-up. Participants agreed that a meaningful outcome of learning would be guidance for practical systems to routinely provide information for quality improvement and for the delivery of follow-up information and care to women and newborns.

Two topics were considered nearly equally important: “Use of tools such as clinical checklists and audits for cesarean delivery (CD) and peripartum hysterectomy to enhance decision-making as part of quality improvement” and “Trial of labor after CD (TOLAC) or vaginal birth after CD (VBAC): LMIC practices, availability/coverage, outcomes.” Participants considered that tools for clinical decision-making could have great potential for impact on quality and outcomes of care within efforts to optimize the CD rate and strengthen quality improvement systems. However, given the availability of many tools, learning on proper implementation and consistent use was deemed more important than the development and documentation of a particular tool. The importance of learning on TOLAC and VBAC was emphasized despite the challenges inherent in its learning (e.g., limited provision). Participants also discussed the value of linking research on TOLAC and VBAC to activities related to improving use of the Robson classification system.

Discussants considered learning about intrapartum/midwifery care practices to reduce unnecessary CD as important, although at a lower priority than post-surgical follow-up, tools for decision-making, and TOLAC/VBAC. In order to focus this topic on meaningful learning, participants suggested stratifying the topic across two sub-topics: (1) specific, effective practices within intrapartum care (e.g., tools to manage labor and ensure birth accompaniment and respectful care) and (2) ways to strengthen midwifery-led models of care that favor less intervention and empower patients, including by understanding why countries trying to implement such models are still struggling to make progress. Moreover, how these issues vary for midwives who work individually versus within a team was noted as an important contextual and learning design factor.

The ranking consultation and post-consultation synthesis produced the following research and learning topics for surgical obstetric care (table 7):

- Use of post-discharge/postoperative visits (in-person or telehealth) to monitor postoperative morbidity and neonatal outcomes
- Use of tools, such as clinical checklists and audits for CD and peripartum hysterectomy, to enhance decision-making as part of quality improvement
- TOLAC or VBAC: LMIC practices, availability/coverage, and outcomes
- Intrapartum/midwifery care practices to reduce unnecessary CD, specifically: (1) effective practices within intrapartum care (e.g., tools to manage labor and ensure birth accompaniment and respectful care) and (2) ways to strengthen midwifery-led models of care that favor less intervention and empower patients, including by understanding barriers to progress

### Fistula Prevention and Treatment

#### Rating

Of the 20 fistula topics rated, six rated in the top 25% and were presented back for ranking (table 4). Among these, the topic with the highest overall rating was “High-impact social and behavior change strategies for community engagement for fistula prevention” (table 4).

**Table 4.**
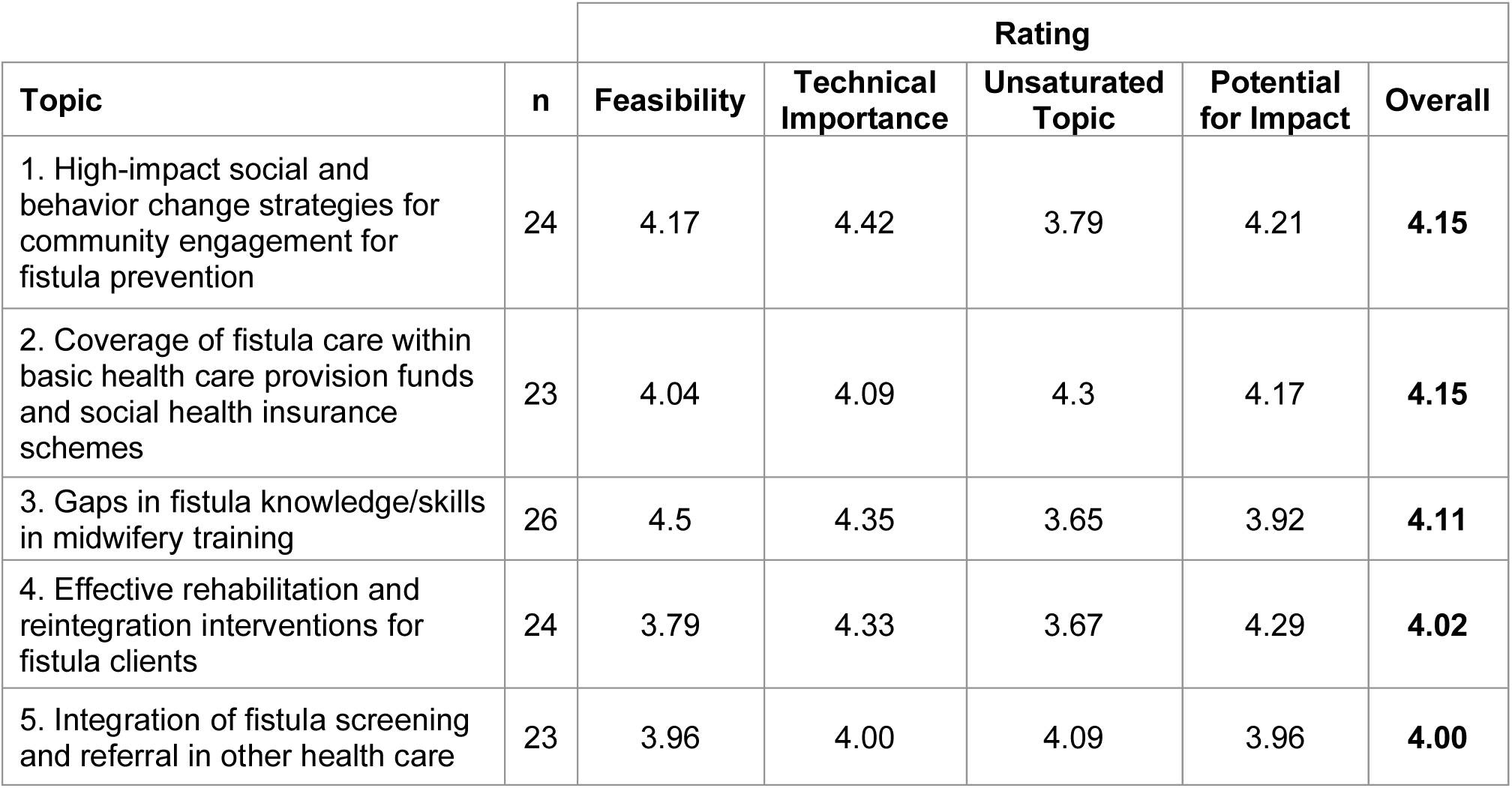
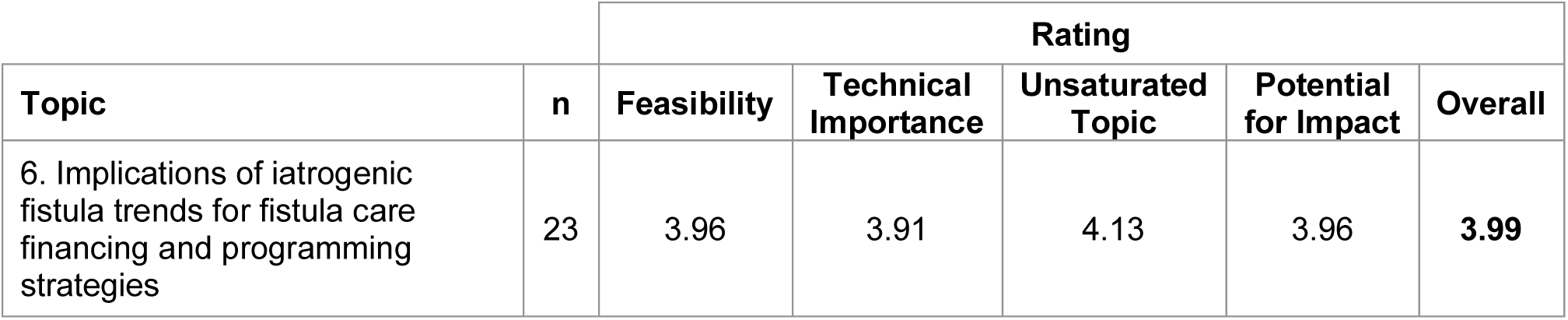
Fistula Prevention and Treatment Topics Ranked for Prioritization by the Expert Group.

| Topic | n | Rating |  |  |  |  |
| --- | --- | --- | --- | --- | --- | --- |
|  |  | Feasibility | Technical Importance | Unsaturated Topic | Potential for Impact | Overall |
| 1. High-impact social and behavior change strategies for community engagement for fistula prevention | 24 | 4.17 | 4.42 | 3.79 | 4.21 | **4.15** |
| 2. Coverage of fistula care within basic health care provision funds and social health insurance schemes | 23 | 4.04 | 4.09 | 4.3 | 4.17 | **4.15** |
| 3. Gaps in fistula knowledge/skills in midwifery training | 26 | 4.5 | 4.35 | 3.65 | 3.92 | **4.11** |
| 4. Effective rehabilitation and reintegration interventions for fistula clients | 24 | 3.79 | 4.33 | 3.67 | 4.29 | **4.02** |
| 5. Integration of fistula screening and referral in other health care | 23 | 3.96 | 4.00 | 4.09 | 3.96 | **4.00** |
|  |  | Feasibility | Technical Importance | Unsaturated Topic | Potential for Impact | Overall |
| 6. Implications of iatrogenic fistula trends for fistula care financing and programming strategies | 23 | 3.96 | 3.91 | 4.13 | 3.96 | **3.99** |

#### Ranking and Synthesis

The group initially identified three highest-rated topics: social behavior change (SBC), coverage/financing, gaps in knowledge/skills (table 4), but did not want to rank them relative to one another as they were viewed as complementary. Participants noted the importance of learning regarding strengthening provider skills, including among midwives, to prevent fistula through appropriate management or referral of prolonged/obstructed labor, but also noted that such capacity strengthening may not achieve meaningful impact if patients cannot afford or access services. Additionally, there will be no demand for care or behavior change if community members are unaware of service availability, fistula risk factors, and behaviors that can prevent fistula.

When encouraged to rank the topics, “Gaps in fistula knowledge/skills in midwifery training” emerged as the highest priority. However, participants recommended that the topic be expanded to focus on provider competency more broadly. They suggested this topic include exploring where new cases are coming from, technical skills for CD and fistula repair, and rehabilitation and reintegration training and care availability. Moreover, participants noted that this topic should address the full continuum from the community to the facility to the infrastructure of the health system. In addition, participants interpreted the topic of fistula “care” broadly, potentially including prevention and treatment, screening and referral, and rehabilitation services.

Although three other topics (rehabilitation and reintegration, integration of screening and referral into other services, and iatrogenic fistula) were considered important (table 4), the group concluded that they could be integrated into the remaining topics. Through these discussions, the group agreed on the following three priority topics (table 7):

- Gaps in the knowledge/skills required for provision of holistic fistula care, including midwifery/obstetric care, surgical and nonsurgical fistula repair, and rehabilitation
- High-impact SBC strategies for community engagement for fistula prevention and care
- Coverage of fistula care within basic healthcare provision funds and social health insurance schemes, including prevention, treatment, screening and referral, and rehabilitation care

### Family Planning

#### Rating

Of the eight topics rated by the expert group, two topics rated in the top 25%; given this small number, the top five were presented back for ranking for parity with the other technical areas (table 5). Among these, the topic with the highest overall rating was “Task sharing for implant removal procedures: feasibility, effectiveness, recommended settings” (table 5).

**Table 5.** Family Planning Topics Ranked for Prioritization by the Expert Group.

| Topic | n | Rating |  |  |  |  |
| --- | --- | --- | --- | --- | --- | --- |
|  |  | Feasibility | Technical Importance | Unsaturated Topic | Potential for Impact | Overall |
| 1. Task sharing for implant removal procedures: feasibility, effectiveness, recommended settings | 24 | 4.38 | 4.13 | 3.71 | 4.17 | **4.09** |
| 2. Effectiveness of interventions to increase post-abortion contraception uptake and continuance | 24 | 4.08 | 4.29 | 3.08 | 4.46 | **3.98** |
| 3. SBC strategies focusing on men as supportive partners, co-decision-makers, and clients of LARCs/PMs | 24 | 4.04 | 3.96 | 3.50 | 3.96 | **3.86** |
| 4. Impact of human-centered designed social behavior change (SBC) activities on long-acting reversible contraceptives / permanent methods (LARC/PM) awareness and uptake | 25 | 3.84 | 4.20 | 3.29 | 4.04 | **3.84** |
|  |  | Feasibility | Technical Importance | Unsaturated Topic | Potential for Impact | Overall |
| 5. Effective strategies for youth-led organizations to increase capacity to implement evidence-informed interventions to expand access to LARCs/PMs | 24 | 3.58 | 3.96 | 3.67 | 4.00 | **3.80** |

#### Ranking and Synthesis

When reviewing the top-rated family planning topics (table 5), the small group discussed the need for some topics to be further expanded. For example, participants felt that the postabortion contraception uptake and continuance topic warranted expansion to include postpartum family planning and the task sharing for implant removal topic needed to include intrauterine device removal. The group proposed combining the SBC topics due to significant overlap. Although the group recognized the merit of pursuing learning on strategies for youth-led organizations to increase capacity vis-à-vis expanding access to LARCs, the inappropriateness of PMs for that age group was discussed, and the topic was narrowed to life course counseling, consent, and/or policies for PMs. However, the group discussed other research and implementing partners that are already engaged on this topic and recommended dropping it from consideration in this exercise.

The group also discussed topics that were not highly rated in the survey but considered important, nonetheless. Topics related to vasectomy advocacy were noted both due to its underrepresentation in research efforts and its implications for gender equity in family planning. Participants noted that learning related to ensuring matching of supply and demand for vasectomy could be especially meaningful given the method’s historical challenges. The group also discussed the need for learning on tools to support surgical safety and quality within surgical family planning methods, including checklists and monitoring postoperative outcomes. The final list of topics for the family planning group was (table 7):

- Task sharing for LARC removal procedures: feasibility, effectiveness, and recommended settings
- Effectiveness of interventions to increase voluntary postabortion and postpartum contraception—including LARCs and permanent methods (PMs)—uptake and continuance
- LARC/PM SBC strategies, including impact of human-centered design activities to strengthen awareness and uptake, and strategies focusing on men as supportive partners, co-decision-makers, and clients

### Cross-Cutting Safe Surgery

#### Rating

Five of the 17 cross cutting safe surgery topics rated by the expert group were in the top 25% and were presented back for ranking (table 6). Among these, the topic with the highest overall rating was “Effective strategies to strengthen key aspects of the safe surgery ecosystem (e.g., anesthesia supplies, blood, and oxygen)” (table 6).

**Table 6.**
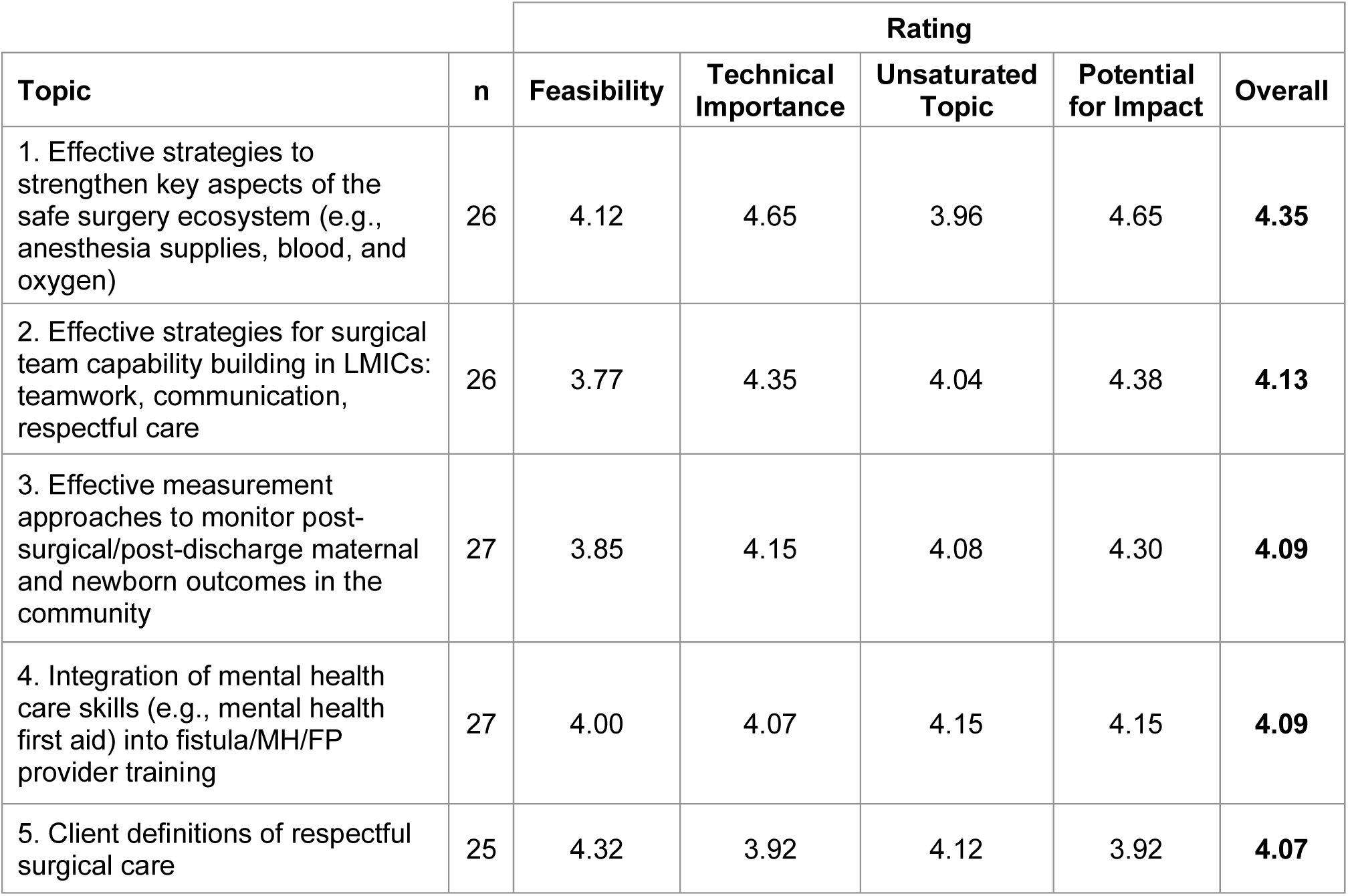
Cross-Cutting Safe Surgery Topics Ranked for Prioritization by the Expert Group.

| Topic | n | Rating |  |  |  |  |
| --- | --- | --- | --- | --- | --- | --- |
|  |  | Feasibility | Technical Importance | Unsaturated Topic | Potential for Impact | Overall |
| 1. Effective strategies to strengthen key aspects of the safe surgery ecosystem (e.g., anesthesia supplies, blood, and oxygen) | 26 | 4.12 | 4.65 | 3.96 | 4.65 | **4.35** |
| 2. Effective strategies for surgical team capability building in LMICs: teamwork, communication, respectful care | 26 | 3.77 | 4.35 | 4.04 | 4.38 | **4.13** |
| 3. Effective measurement approaches to monitor post-surgical/post-discharge maternal and newborn outcomes in the community | 27 | 3.85 | 4.15 | 4.08 | 4.30 | **4.09** |
| 4. Integration of mental health care skills (e.g., mental health first aid) into fistula/MH/FP provider training | 27 | 4.00 | 4.07 | 4.15 | 4.15 | **4.09** |
| 5. Client definitions of respectful surgical care | 25 | 4.32 | 3.92 | 4.12 | 3.92 | **4.07** |

#### Ranking and Synthesis

Many of the participants thought that all the topics were important and agreed with their inclusion in the ranking process. However, participants felt that ranking the topics was difficult and contextual; priorities may differ across geography, for example. Additionally, some participants thought that achieving impact on the respectful care topic could be challenging as respectful maternity care more broadly has been incorporated into many policies and procedures already but is still often poorly operationalized in routine maternal care.

Participants agreed that evidence on post-discharge maternal and newborn outcomes at the community-level is limited and that learning on these outcomes will address gaps in the quality of care and service delivery processes. They emphasized that the post-discharge period should be a critical part of overall quality improvement initiatives. The group agreed upon the following ranking (table 7):

- Effective measurement approaches to monitor postsurgical/post-discharge maternal and newborn outcomes in the community
- Effective strategies to strengthen key aspects of the safe surgery ecosystem (e.g., anesthesia supplies, blood, and oxygen)
- Effective strategies for surgical team capability building in LMICs, including teamwork, communication, and respectful care
- Client definitions of respectful surgical care
- Integration of mental healthcare skills (e.g., mental health first aid) into fistula, MH, and FP provider training to support provision of mental health assessments of and care to patients

### Summary of Prioritized Topics

Following one round of rating and one round of ranking, a total of 15 topics were prioritized across the four technical areas explored in this consultation (table 7).

**Table 7.**
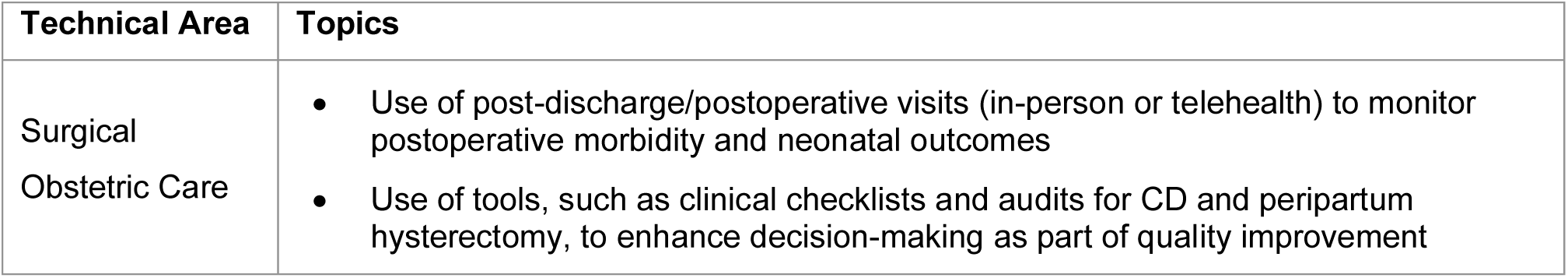
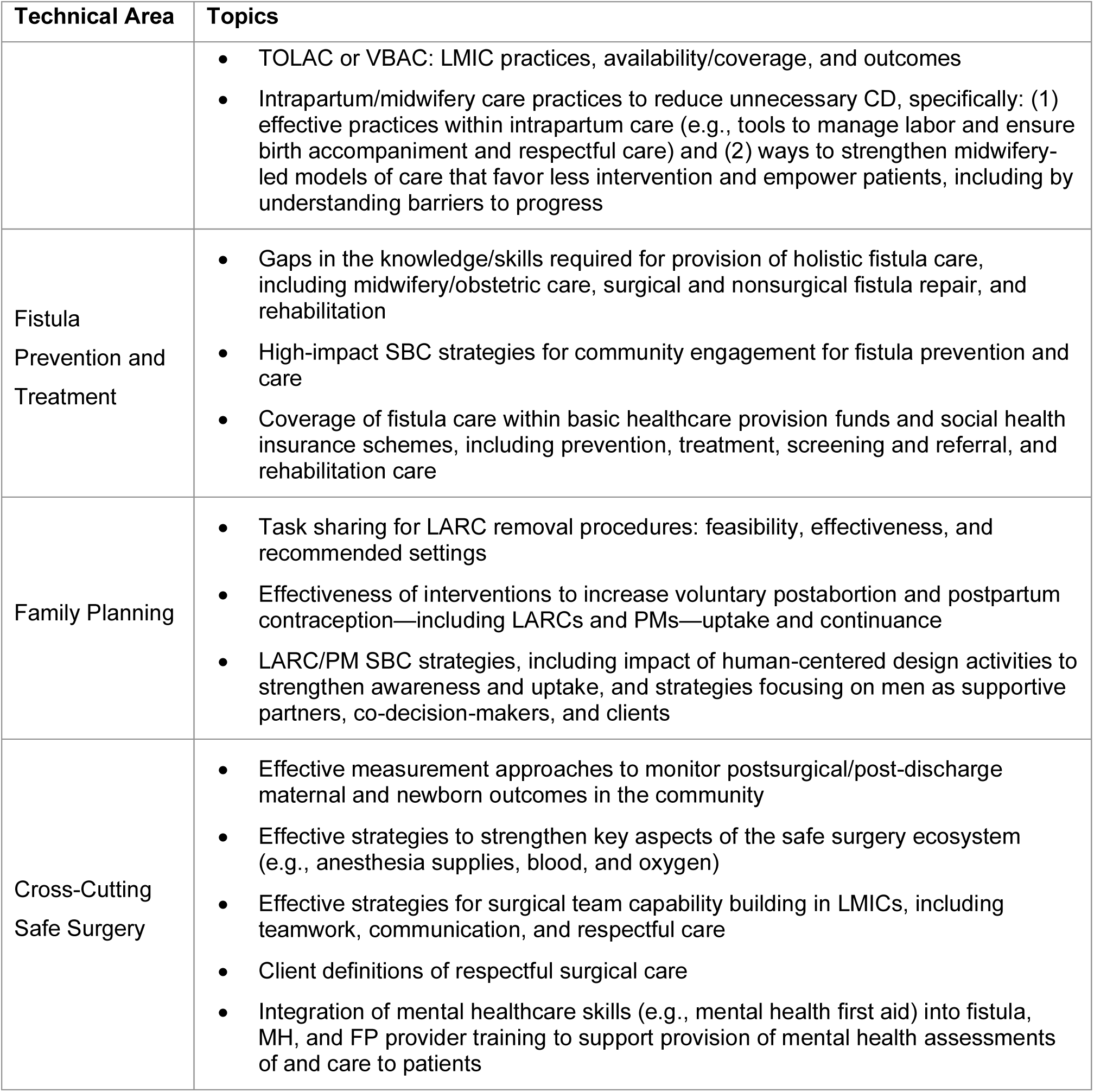
List of Topics Prioritized for Research and Learning.

## Discussion

### Summary of findings

This expert consultation, comprised of a rating survey and a ranking exercise, yielded a core set of actionable research and learning topics across four technical areas: surgical obstetric care, fistula prevention and treatment, family planning, and cross-cutting safe surgery. Although the topics were divided into four distinct categories, this exercise demonstrated the interdependence of topics in safe surgery across technical areas. The prioritization of topics on TOLAC/VBAC and intrapartum/midwifery practices to reduce unnecessary CD not only align with WHO recommendations on performing CD only when medically indicated,^31^ they are also linked to the prevention of iatrogenic fistulas from CD complications. SBC strategies were recognized as important for both fistula prevention and treatment and surgical family planning. Many topics touched on the perioperative period (capacity building through teamwork, use of tools for decision-making as part of quality improvement, blood/anesthesia/oxygen, task sharing for LARC removal procedures). Two topics, one each in surgical obstetric care and cross-cutting safe surgery, specifically called out the postoperative period as highly neglected in both evidence and practice. Several domains of respectful maternity care are represented in the prioritized topics across the four technical areas.^32^

### Strengths and limitations

This process adapted an approach and criteria previously developed and applied by a comparable global project to generate a focused priority learning agenda and develop a research portfolio targeting the identified priorities.^11^ The experience and strong engagement of the expert group, comprehensive rating criteria, and iterative nature of the process represent strengths of this consultation. The group of international experts held expertise spanning research and evaluation, clinical practice, and development assistance and implementation across the four focus technical areas. The rating criteria enabled the identification of topics where need for further investigation is greatest, yet whose investigation could be done practically in the LMIC context. Moreover, the criteria definitions gained additional precision from discussions with the expert group; for example, recommendations resulted in sustainability being included in the criteria definition of potential for impact. The use of a virtual format enabled greater representation of LMIC participants and avoided travel-related emissions. The process enabled expeditious prioritization, enabling the project to focus on pursuing research activities and generating evidence.

However, the approach utilized also featured several limitations. First, as implemented, the representation of the expert group was limited with respect to affiliation. Representation was largely from international organizations and four African universities; no Ministry of Health officials participated, nor did women, families, or communities. Second, we did not collect data on gender, geographic location, or occupation (including experience performing focal surgical procedures). Third, although 39 individuals engaged in the rating survey, some attrition was observed (e.g., only 23 responses to the fistula topics), and fewer than 10 people engaged in each of the technical area-specific small group discussions. Fourth, the family planning topics emerging from this process (as designed and implemented) did not generate priorities on voluntary surgical contraceptive methods. It is possible that any of these limitations, including the small group composition, could have introduced bias, especially at the rating stage where small changes in rating could have resulted in any given topic being included or excluded from discussion during the ranking stage. We acknowledge that the composition of topics (both included initially and prioritized) could have been different if any of these design factors were altered (e.g., foregoing a discussion group-based model in favor of a survey-based model that could have accommodated many more participants).

These findings should be interpreted, therefore, as research priorities for a project working in LMIC, not an agenda comprehensively applying to all LMIC. As the surgical obstetrics and family planning evidence base continues to build, we also acknowledge that new lines of inquiry will emerge, and research topics will need to be re-prioritized. Several lessons from this process can inform future prioritization efforts (Box 2).

In addition to the aforementioned limitations, we believe that the deficit of topics on voluntary surgical contraceptive methods reflects an overall lack of attention paid to permanent family planning methods, overall. That said, we intentionally set the scope of the prioritization to cover LARCs and PMs both because these are the project’s technical areas of focus and because they are procedure-based methods using surgical instruments. However, the topic we included in the initial list on trends and enabling factors in global vasectomy availability and uptake was not rated highly enough to warrant discussion during the ranking stage. This could have been because the expert group rating topics was multidisciplinary in nature and was mandated with approximating priorities in the *overall* landscape of surgical obstetrics and family planning. Had the rating process been restricted to participants exclusively involved in the family planning community of practice, it is possible that this topic could have been more highly rated. It is also important to note that a recently published review sheds some light on global vasectomy trends and enabling factors in uptake, as well as persistent barriers to this method receiving advocacy and attention,^33^ and that voluntary surgical contraceptive programming can be studied through the lens of the cross-cutting safe surgery topics prioritized through this process. We nevertheless acknowledge that our approaches could have been adjusted to increase the curation of topics relating to vasectomy and tubal ligation.

#### Box 2.

##### Lessons Learned for Future Prioritization Efforts

- Time-limited projects seeking to generate their research and learning agendas through prioritization methods must balance the time consumed by generating consensus-based priorities with the time needed to conduct studies to generate evidence.
- Consultation-based methods can generate rich and contextualized insights, which is helpful for developing research priorities in broad, interdisciplinary spaces (such as safe surgery). However, such methods are inherently limited in the number of participants they can accommodate.
- Developing research priorities that comprehensively apply to all LMIC will require methods that can engage a wider array of actors (e.g., Ministry of Health officials, civil society organizations, academics based in LMICs, patients). Survey-based consensus-building methods (e.g., CHNRI) may be more appropriate for this objective.
- Future prioritization processes ought to gather participant data on occupation (including experience performing surgery, by procedure), gender, and geographic location so that analyses can be done to determine whether the research priorities generated reflects the priorities of key subgroups.

### Implications for research and practice

It is impossible to avert a significant proportion of maternal morbidity and mortality in LMIC without access to high-quality surgical services delivered to the right patient at the right time. In many parts of the world, however, access to surgery is limited,^3^ referral systems for emergency obstetric conditions remain weak,^34^ preventable peri-operative complications persist,^6,35^ and post-operative care needs reinforcement.^36^ From a human resources for health standpoint, increasing the density of surgeons, obstetricians, and anesthesiologists is correlated with a decline in the maternal mortality ratio, especially in settings with less than twenty specialists per 100,000 population.^2,37^ Where it is not possible to easily scale up the number of specialists, surgical task sharing to non-specialist physicians and non-physician clinicians has been implemented for a variety of procedures in many settings.^38,39^ In parallel, the number of cesarean sections, already the most frequently performed surgical procedure worldwide, has increased in recent years, and is expected to continue increasing.^40^ Therefore, strengthening the provision of safe essential surgery in settings with fragile health systems through obstetric, fistula, and family planning programming can both prevent maternal and newborn morbidity and mortality as well as serve as a framework for strengthening the entire surgical ecosystem and the ensemble of surgical services provided by health facilities.

This learning agenda contributes to efforts at reducing maternal morbidity and mortality through programmatic and clinical learning across the safe surgery ecosystem, which encompasses processes involved in seeking, reaching, and receiving quality surgical care as well as the linkages after surgical care. High impact SBC strategies for LARC/PM and fistula prevention and care, TOLAC/VBAC, and enhancing intrapartum/midwifery care practices to reduce unnecessary CD represent learning priorities that aim to prevent complications necessitating surgical care or improve women’s seeking of surgical care when needed. Several topics explore the provision of surgical care, including strengthening the perioperative environment (e.g., anesthesia supplies, blood, oxygen), surgical team capacity building, respectful surgical care, tools for strengthening decision-making, and task sharing for LARC removal procedures. Lastly, several topics touch upon strengthening continuity of care after surgery were prioritized, including the use of post-discharge visits, monitoring of post-operative outcomes, integration of mental healthcare skills in surgical procedures, and gaps in the provision of holistic fistula care. Referral for surgical and emergency obstetric and newborn care is underrepresented in this agenda, however, a significant amount of research has been done on this topic and a recently published systematic review of referral interventions for obstetric emergencies proposes a logic model for “studying, understanding and improving” such processes.^34^

Because the research priorities emerging from this process are programmatic and clinical in nature, this agenda neglects topics that may be a few steps removed from service delivery but may nevertheless, be crucial for strengthening surgical systems in the long-term. For example, this agenda does not cover policy questions, such as the generation of political support for safe surgery,^41^ or effective strategies for developing and implementing National Surgical Obstetric and Anesthesia plans in more LMIC. This outcome, though unintended, may be the result of strong representation of implementing partners in developing this agenda and because the rating criteria, specifically feasibility and potential for impact, may have contributed to prioritization of topics whose pursuit could achieve the most impact with the fewest resources. However, this agenda is not intended to stand alone, but rather to complement other existing learning agendas at the global and national levels, including on maternal health, family planning, LMIC safe surgery (e.g., general, pediatrics, orthopedics), and health systems and policy research.

### What’s next? Using the agenda to guide the implementation of an integrated surgical obstetrics and family planning project’s learning activities

The MOMENTUM Safe Surgery in Family Planning and Obstetrics project is using this learning agenda to guide its research and learning strategy. As such, the project has launched a series of activities involving special studies and targeted literature reviews (Table 8). Whereas some of these activities were initiated at the global level, others were launched in response to research priorities established by national and local stakeholders. Examples of the latter include the measurement of post-surgical complications before and after the implementation of a peripartum surgery diploma in Mali and the Robson classification study in Senegal (Table 8). One of the benefits of the broad nature of many of the learning agenda topics is that they can be readily aligned with research priorities established at the national and subnational levels. Although this agenda has guided the project’s research and learning efforts, we have also supported Ministries of Health in generating evidence for topics falling outside of the learning agenda’s scope. As projects consider this type of prioritization methodology for developing their learning agendas, the research they support ought to align with local priorities.

**Table 8.**
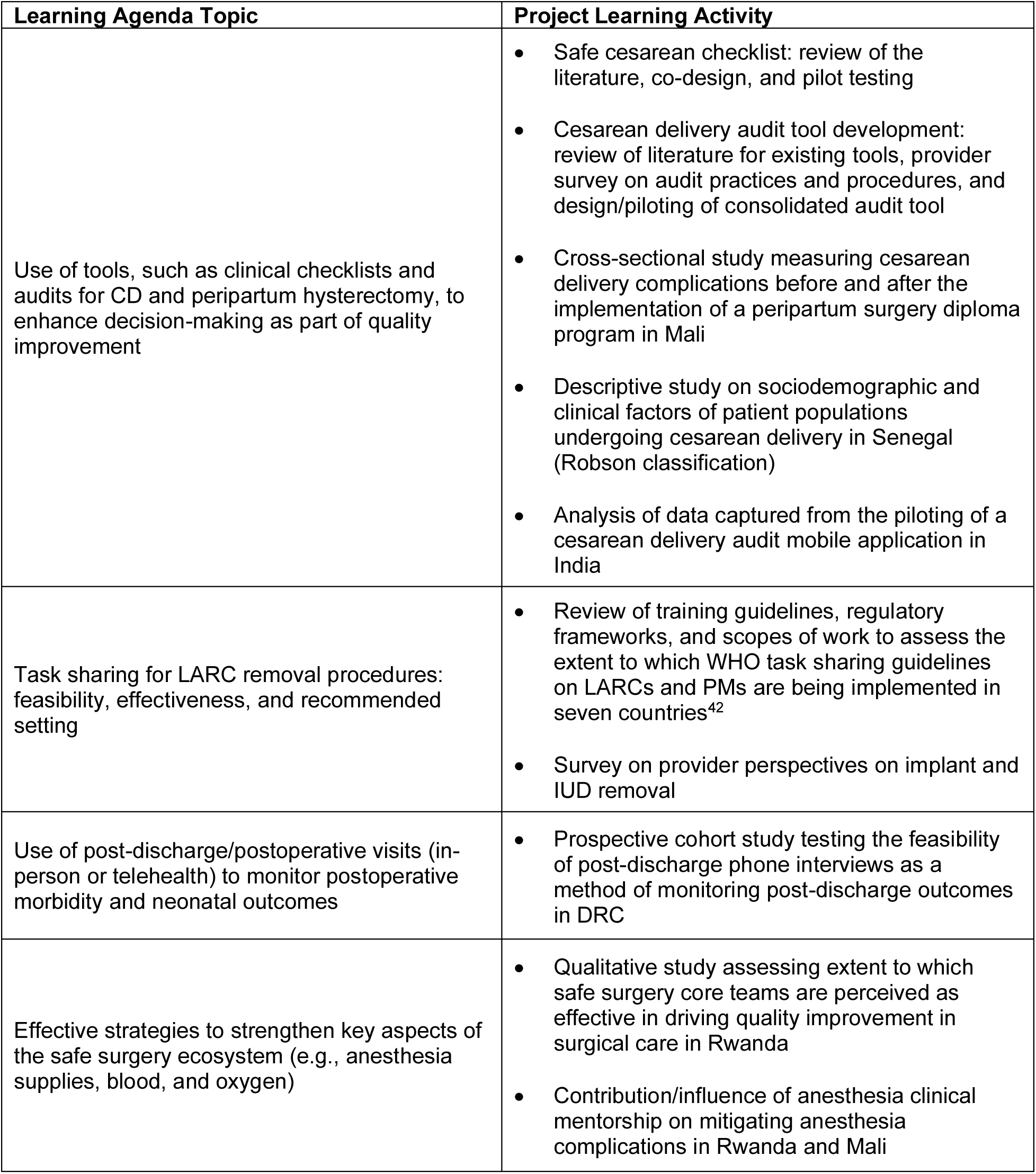
Illustrative examples of project activities contributing to the advancement of learning agenda topics.

| Learning Agenda Topic | Project Learning Activity |
| --- | --- |
| Use of tools, such as clinical checklists and audits for CD and peripartum hysterectomy, to enhance decision-making as part of quality improvement | • Safe cesarean checklist: review of the literature, co-design, and pilot testing<br>• Cesarean delivery audit tool development: review of literature for existing tools, provider survey on audit practices and procedures, and design/piloting of consolidated audit tool<br>• Cross-sectional study measuring cesarean delivery complications before and after the implementation of a peripartum surgery diploma program in Mali<br>• Descriptive study on sociodemographic and clinical factors of patient populations undergoing cesarean delivery in Senegal (Robson classification)<br>• Analysis of data captured from the piloting of a cesarean delivery audit mobile application in India |
| Task sharing for LARC removal procedures: feasibility, effectiveness, and recommended setting | • Review of training guidelines, regulatory frameworks, and scopes of work to assess the extent to which WHO task sharing guidelines on LARCs and PMs are being implemented in seven countries<sup>42</sup><br>• Survey on provider perspectives on implant and IUD removal |
| Use of post-discharge/postoperative visits (in-person or telehealth) to monitor postoperative morbidity and neonatal outcomes | • Prospective cohort study testing the feasibility of post-discharge phone interviews as a method of monitoring post-discharge outcomes in DRC |
| Effective strategies to strengthen key aspects of the safe surgery ecosystem (e.g., anesthesia supplies, blood, and oxygen) | • Qualitative study assessing extent to which safe surgery core teams are perceived as effective in driving quality improvement in surgical care in Rwanda<br>• Contribution/influence of anesthesia clinical mentorship on mitigating anesthesia complications in Rwanda and Mali |

### How can the broader maternal health, family planning, and health systems communities of practice contribute to advancing these priority research and learning topics?

Although this agenda was primarily developed for a global project working in LMICs, any one time-bound, donor-funded project can only take on a limited level of learning on such a diverse set of priorities. Evidence on the topics prioritized in this collaborative learning agenda can be advanced by domestic and donor agencies, research institutions, and implementing partners supporting maternal and reproductive health service strengthening.

Indeed, priorities at the national and sub-national levels may not necessarily align with priorities at the global level. As such, research prioritization and evidence generation efforts at the national and sub-national levels ought to be led by Ministry of Health officials as well as their partners (e.g. academia, implementing partners, civil society, the private sector, and community-based organizations). As mentioned above, this agenda was primarily developed as a project’s learning agenda and does not intend to substitute national/subnational efforts, but rather to complement them. The topics prioritized in this agenda may, however, support national and sub-national stakeholders in identifying potential lines of inquiry to prioritize or interventions to study.

Although siloed/vertical nature of much development assistance for health presents a challenge for learning resource mobilization given the integrated/horizontal nature of safe surgery systems,^41^ large-scale donor-funded health programs may want to consider adopting these topics when developing their evidence generation strategies. Stratifying this learning exercise by obstetrics, fistula, family planning, and cross-cutting safe surgery topics can enable implementing partners to consider topics that align with their programmatic mandates – whether vertical or integrated. A maternal health program may, for example, consider investigating the obstetrics topics. As basic and essential surgery is increasingly being recognized as a key component of primary health care,^43^ an integrated primary health care program working across several technical areas may consider designing implementation research studies with these topics in mind. Many of the cross-cutting topics identified in this agenda can be studied even through the lens of vertical interventions. For example, client definitions of respectful care or strategies for surgical team capacity building can be studied in the context of cesarean deliveries, permanent family planning methods, and/or fistula repair.

That said, the prioritization of topics in this agenda that inherently build on and overlap with each other, further emphasizes the integrated nature of the common ecosystem that support surgeries across different technical areas (e.g., cross-cadre surgical teams, safe blood and oxygen systems, and monitoring of post-surgical outcomes) and suggest opportunities for funding research and programming that can be conducted and applied to a range of services. Given the relative newness of global safe surgery as a global public health priority, we view the development of this agenda as a starting point, not an end point, for a dialogue across different communities of practices working at different levels (academics, funders, and implementers).

## Conclusion

Through an expert consultation, research and learning topics were identified and prioritized across four technical areas of a safe surgery project that compromise important interventions in maternal health and family planning that are growing in volume and whose availability, access, quality, and safety need to be strengthened for optimal impact and use of resources. These topics may warrant consideration for exploration in the research and learning efforts of national, regional, and global maternal health, and family planning initiatives, including via targeted investment of domestic and donor resources.

## Data Availability

All data produced in the present study are available upon reasonable request to the authors

## Acknowledgements

The authors thank the subject matter experts that participated in the consultations, Lara Vaz and Louise Day for providing advice on methods, and the Maternal and Child Health and Nutrition and Population and Reproductive Health teams at USAID for their reviews of this article.

## Disclaimer

NA

## Supplementary materials

### MOMENTUM Safe Surgery in Family Planning and Obstetrics: Learning Agenda Topics Rating Survey

Thank you for participating in the first consultation to develop the MOMENTUM Safe Surgery in Family Planning and Obstetrics Global Learning Agenda. Following this first meeting on 31 Jan 2022, the learning topics and rating criteria were refined based on your feedback, including incorporating elements of the Child Health and Nutrition Research Initiative (CHNRI) prioritization criteria.^1^ We have also added a number of additional learning topics based on your suggestions. This survey represents the next step in the development of the project’s global learning agenda. We are asking you to rate proposed learning topics according to the following criteria:

#### Feasibility

- Can be addressed with ethical research/learning
- Can be answered within the resources and timeframe available

#### Technical importance

- Has notable gaps in evidence from LMIC settings
- Is of interest, debate, and discussion among technical peers and leaders in the field
- Has the potential for a large effect on RMNCAH outcomes and/or the wellbeing of women and newborns

#### Unsaturated topic

- Extensive, quality research/learning relevant to LMIC settings is not already being conducted
- Avoids redundancy with other projects’ learning agendas (where cross-project learning would not be useful/efficient)

#### Potential for program impact

- Research/learning can result in effective and sustainable interventions/programs
- Aligned with global and national priorities and commitments
- Research/learning can clearly lead to action and change how services or programs are delivered in a short time-frame
- Research/learning has potential to generate guidance for large-scale implementation and sustainability through existing systems
- Prioritizes marginalized groups and/or seeks to reduce inequity in access to services/coverage
- Relevance to multiple LMIC

### Interpreting the learning topics

In this survey, we are asking you to rate topics, rather than precise research questions or specific aims. If a given topic is included in the MOMENTUM Safe Surgery in Family Planning and Obstetrics Global Learning Agenda, it may lead to multiple research/learning efforts. For such efforts, specific research/learning questions will be developed and refined to assess the “what”,“why”, and/or “how” (e.g., pathways to impact or mechanisms explaining impact). Please note that prioritizing a topic does not mean it necessarily requires primary data collection – research/learning may involve secondary data analysis, literature review, or synthesis of program data, for example.

### Survey instructions

Please complete the survey on the following pages by applying the four criteria above to rate each potential topic. Rating follows a 1 to 5 scale: 1 is the lowest rating and 5 represents the highest rating. We will aggregate ratings across respondents to determine the most highly rated topics. <u>We suggest allocating an</u> <u>hour to complete this rating process.</u> If you need to complete the survey in several sittings, please be sure not to hit submit until you have completed each section of the survey. Many thanks for your time.

<u>***Note:***</u> This guidance is also provided to you as an email attachment so that you can refer to the criteria definitions while completing the survey.

1. Rudan I. Setting health research priorities using the CHNRI method: IV. Key conceptual advances. J Glob Health. 2016 Jun;6(1):010501.

### Background information

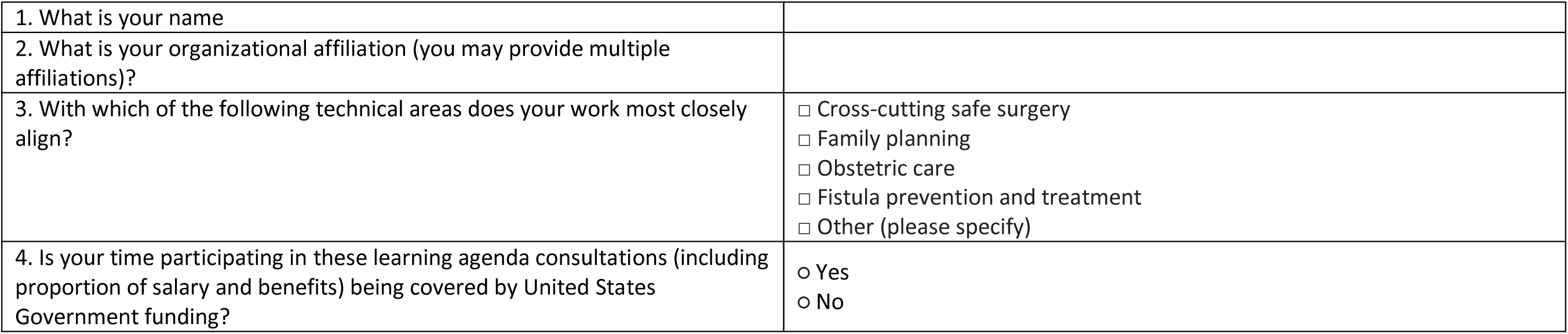

5. Please rate the following topics according to feasibility, technical importance, saturation, and potential for impact (1 is the lowest/poorest rating and 5 is the highest/best rating)

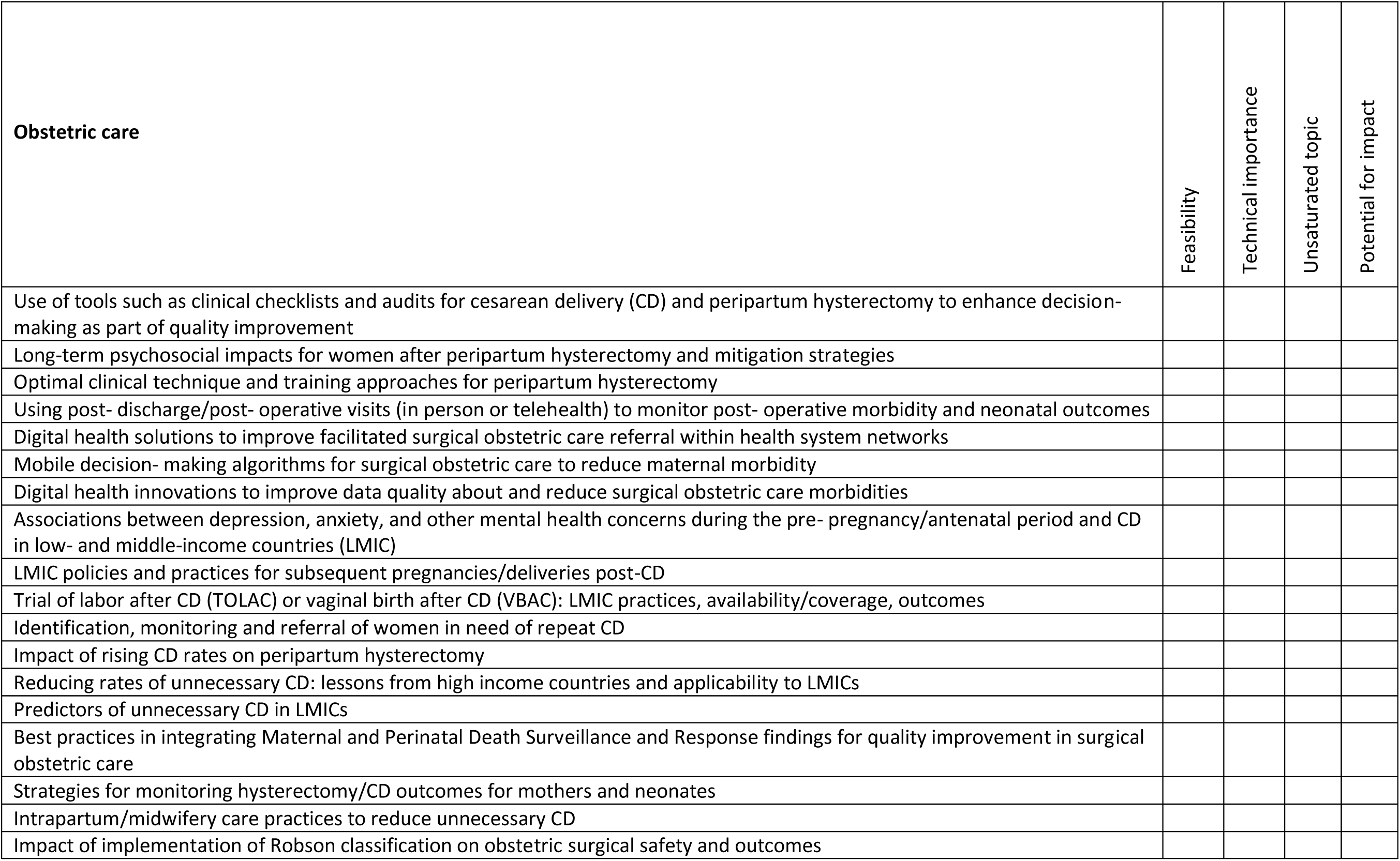

6. Please rate the following topics according to feasibility, technical importance, saturation, and potential for impact (1 is the lowest/poorest rating and 5 is the highest/best rating)

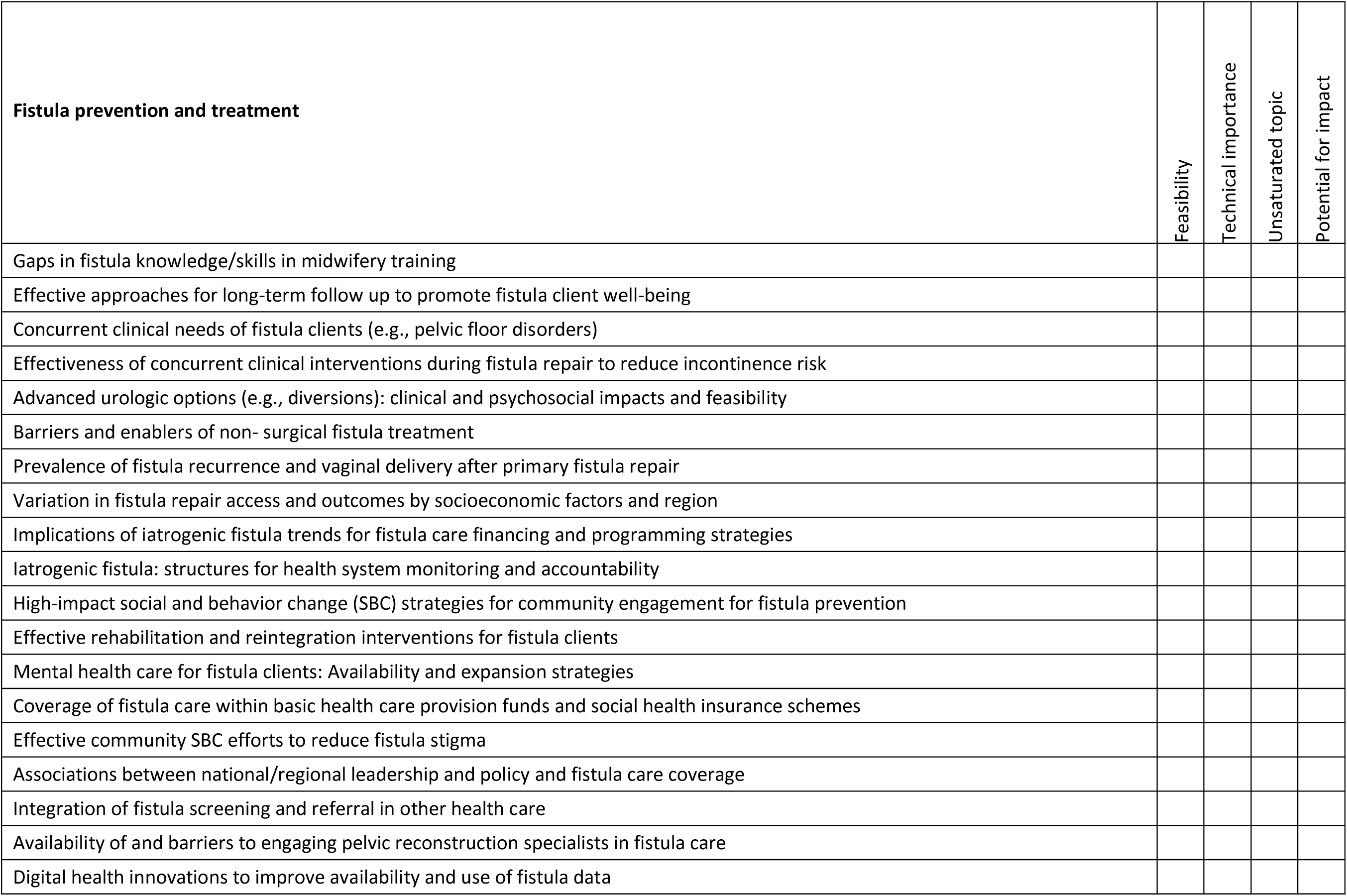
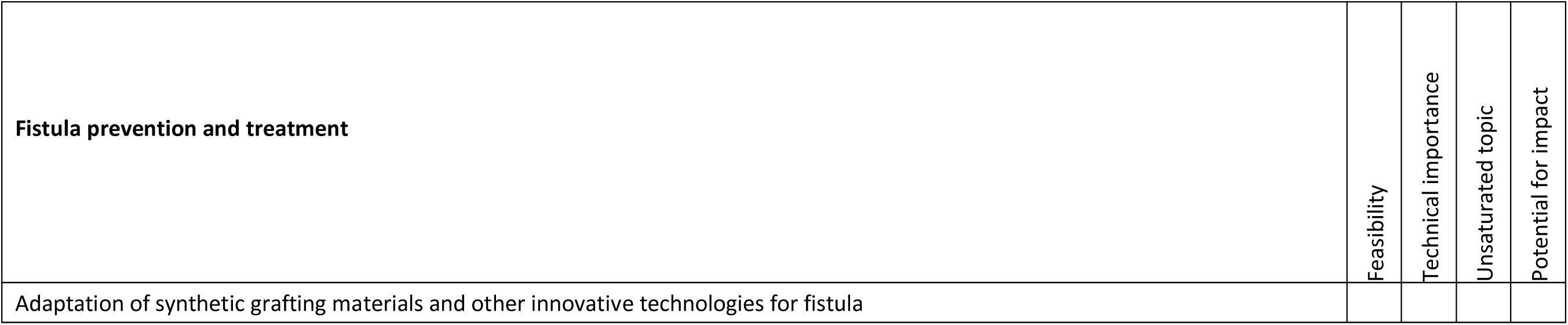

7. Please rate the following topics according to feasibility, technical importance, saturation, and potential for impact (1 is the lowest/poorest rating and 5 is the highest/best rating)

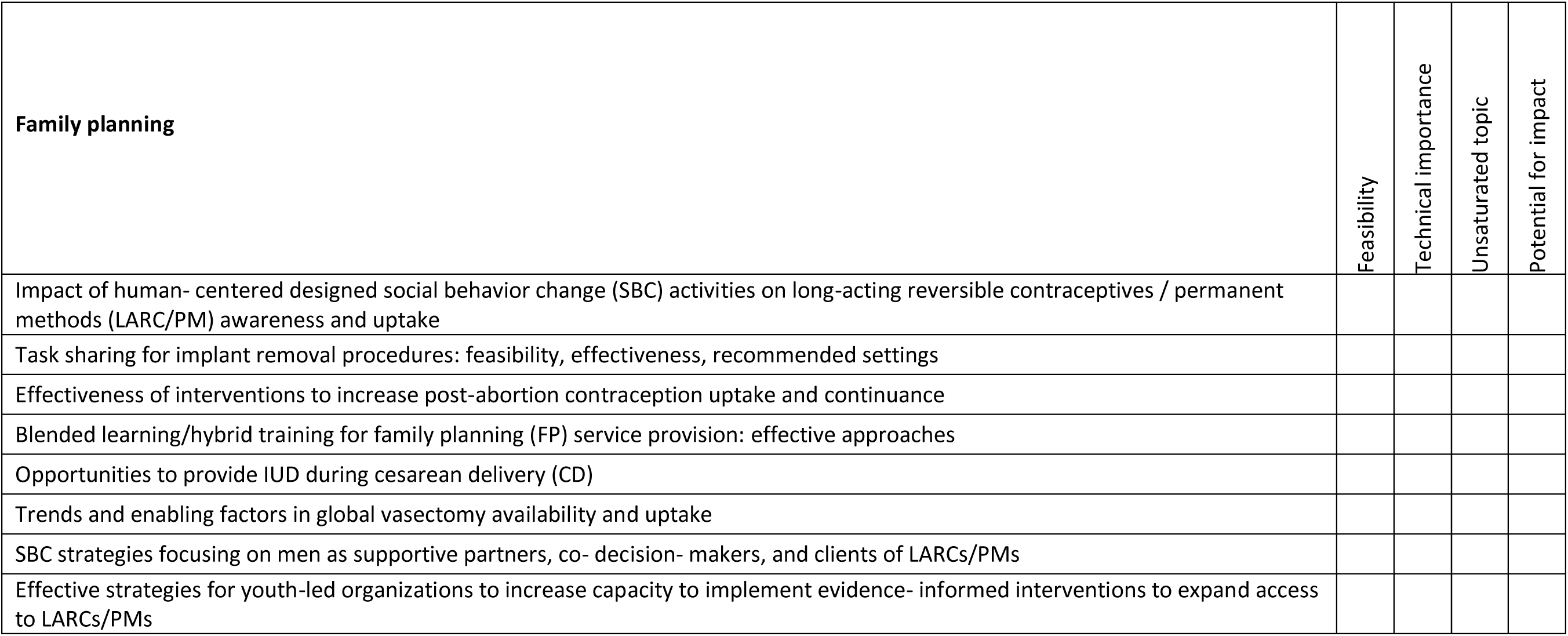

8. Please rate the following topics according to feasibility, technical importance, saturation, and potential for impact (1 is the lowest/poorest rating and 5 is the highest/best rating)

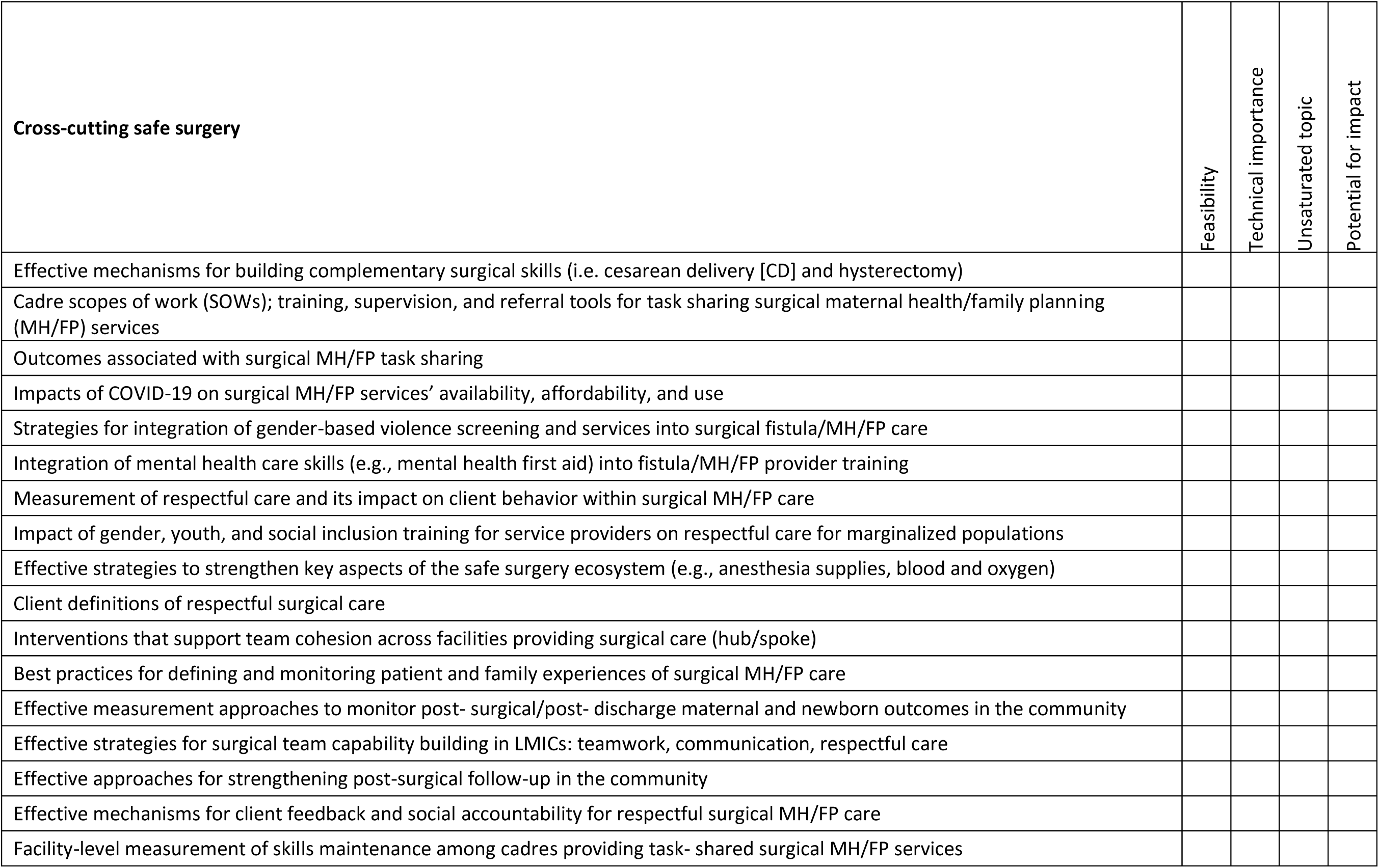

---

**Supplementary Table 1.**
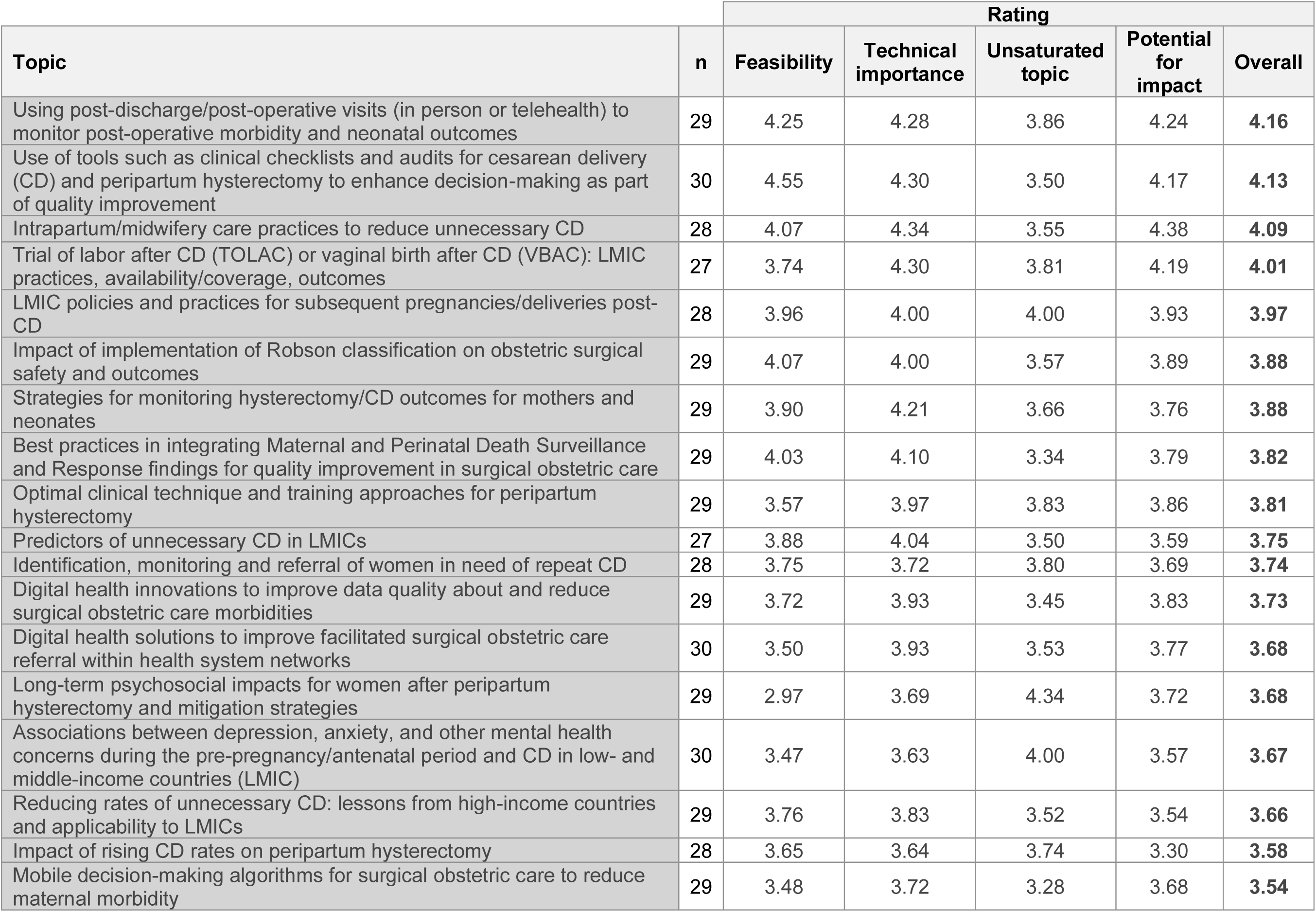
Topics Ratings for Surgical Obstetric Care.

**Supplementary Table 2.** Topics Ratings for Fistula Prevention and Treatment.

| Topic | n | Rating |  |  |  |  |
| --- | --- | --- | --- | --- | --- | --- |
|  |  | Feasibility | Technical importance | Unsaturated topic | Potential for impact | Overall |
| Coverage of fistula care within basic health care provision funds and social health insurance schemes | 23 | 4.04 | 4.09 | 4.30 | 4.17 | **4.15** |
| High-impact social and behavior change (SBC) strategies for community engagement for fistula prevention | 24 | 4.17 | 4.42 | 3.79 | 4.21 | **4.15** |
| Gaps in fistula knowledge/skills in midwifery training | 26 | 4.50 | 4.35 | 3.65 | 3.92 | **4.11** |
| Effective rehabilitation and reintegration interventions for fistula clients | 24 | 3.79 | 4.33 | 3.67 | 4.29 | **4.02** |
| Integration of fistula screening and referral in other health care | 23 | 3.96 | 4.00 | 4.09 | 3.96 | **4.00** |
| Implications of iatrogenic fistula trends for fistula care financing and programming strategies | 23 | 3.96 | 3.91 | 4.13 | 3.96 | **3.99** |
| Mental health care for fistula clients: Availability and expansion strategies | 22 | 3.82 | 3.91 | 4.05 | 4.14 | **3.98** |
| Iatrogenic fistula: structures for health system monitoring and accountability | 23 | 3.70 | 3.91 | 4.22 | 4.00 | **3.96** |
| Effective approaches for long-term follow up to promote fistula client well-being | 26 | 3.77 | 4.15 | 3.85 | 3.92 | **3.92** |
| Effective community SBC efforts to reduce fistula stigma | 23 | 3.91 | 3.96 | 3.52 | 4.09 | **3.87** |
| Barriers and enablers of non-surgical fistula treatment | 24 | 4.17 | 3.79 | 3.83 | 3.57 | **3.84** |
| Effectiveness of concurrent clinical interventions during fistula repair to reduce incontinence risk | 23 | 3.65 | 4.04 | 3.61 | 3.65 | **3.74** |
| Concurrent clinical needs of fistula clients (e.g., pelvic floor disorders) | 23 | 3.87 | 3.87 | 3.48 | 3.61 | **3.71** |
| Availability of and barriers to engaging pelvic reconstruction specialists in fistula care | 22 | 3.59 | 3.59 | 3.68 | 3.41 | **3.57** |
| Variation in fistula repair access and outcomes by socioeconomic factors and region | 23 | 3.78 | 3.74 | 3.26 | 3.39 | **3.54** |
| Digital health innovations to improve availability and use of fistula data | 23 | 3.78 | 3.26 | 3.48 | 3.30 | **3.46** |
| Advanced urologic options (e.g., diversions): clinical and psychosocial impacts and feasibility | 22 | 3.27 | 3.73 | 3.55 | 3.23 | **3.44** |
| Prevalence of fistula recurrence and vaginal delivery after primary fistula repair | 22 | 3.32 | 3.36 | 3.73 | 3.18 | **3.40** |
| Adaptation of synthetic grafting materials and other innovative technologies for fistula | 22 | 2.82 | 3.55 | 3.73 | 3.41 | **3.38** |
| Associations between national/regional leadership and policy and fistula care coverage | 23 | 3.30 | 3.13 | 3.52 | 3.04 | **3.25** |

**Supplementary Table 3.** Topics Ratings for Family Planning.

| Topic | n | Rating |  |  |  |  |
| --- | --- | --- | --- | --- | --- | --- |
|  |  | Feasibility | Technical importance | Unsaturated topic | Potential for impact | Overall |
| Task sharing for implant removal procedures: feasibility, effectiveness, recommended settings | 24 | 4.38 | 4.13 | 3.71 | 4.17 | **4.09** |
| Effectiveness of interventions to increase post-abortion contraception uptake and continuance | 24 | 4.08 | 4.29 | 3.08 | 4.46 | **3.98** |
| SBC strategies focusing on men as supportive partners, co-decision-makers, and clients of LARCs/PMs | 24 | 4.04 | 3.96 | 3.50 | 3.96 | **3.86** |
| Impact of human-centered designed social behavior change (SBC) activities on long-acting reversible contraceptives / permanent methods (LARC/PM) awareness and uptake | 25 | 3.84 | 4.20 | 3.29 | 4.04 | **3.84** |
| Effective strategies for youth-led organizations to increase capacity to implement evidence-informed interventions to expand access to LARCs/PMs | 24 | 3.58 | 3.96 | 3.67 | 4.00 | **3.80** |
| Trends and enabling factors in global vasectomy availability and uptake | 24 | 4.17 | 3.79 | 3.63 | 3.46 | **3.76** |
| Blended learning/hybrid training for family planning (FP) service provision: effective approaches | 24 | 4.04 | 3.83 | 3.25 | 3.83 | **3.74** |
| Opportunities to provide IUD during cesarean delivery (CD) | 24 | 3.79 | 3.58 | 3.88 | 3.63 | **3.72** |

**Supplementary Table 4.** Topics Ratings for Cross-cutting Safe Surgery.

| Topic | n | Rating |  |  |  |  |
| --- | --- | --- | --- | --- | --- | --- |
|  |  | Feasibility | Technical importance | Unsaturated topic | Potential for impact | Overall |
| Effective strategies to strengthen key aspects of the safe surgery ecosystem (e.g., anesthesia supplies, blood and oxygen) | 26 | 4.12 | 4.65 | 3.96 | 4.65 | **4.35** |
| Effective strategies for surgical team capability building in LMICs: teamwork, communication, respectful care | 26 | 3.77 | 4.35 | 4.04 | 4.38 | **4.13** |
| Effective measurement approaches to monitor post-surgical/post-discharge maternal and newborn outcomes in the community | 27 | 3.85 | 4.15 | 4.08 | 4.30 | **4.09** |
| Integration of mental health care skills (e.g., mental health first aid) into fistula/MH/FP provider training | 27 | 4.00 | 4.07 | 4.15 | 4.15 | **4.09** |
| Client definitions of respectful surgical care | 25 | 4.32 | 3.92 | 4.12 | 3.92 | **4.07** |
| Interventions that support team cohesion across facilities providing surgical care (hub/spoke) | 25 | 3.84 | 4.12 | 3.96 | 4.16 | **4.02** |
| Outcomes associated with surgical MH/FP task sharing | 27 | 3.96 | 4.22 | 3.58 | 4.11 | **3.97** |
| Effective mechanisms for building complementary surgical skills (i.e. cesarean delivery [CD] and hysterectomy) | 26 | 3.65 | 4.15 | 3.88 | 4.12 | **3.95** |
| Measurement of respectful care and its impact on client behavior within surgical MH/FP care | 27 | 3.59 | 4.11 | 3.81 | 4.22 | **3.94** |
| Strategies for integration of gender-based violence screening and services into surgical fistula/MH/FP care | 27 | 4.11 | 3.85 | 3.93 | 3.78 | **3.92** |
| Cadre scopes of work (SOWs); training, supervision, and referral tools for task sharing surgical maternal health/family planning (MH/FP) services | 26 | 3.88 | 4.15 | 3.42 | 4.12 | **3.89** |
| Facility-level measurement of skills maintenance among cadres providing task-shared surgical MH/FP services | 26 | 3.88 | 3.92 | 3.77 | 4.00 | **3.89** |
| Impact of gender, youth, and social inclusion training for service providers on respectful care for marginalized populations | 26 | 3.58 | 3.96 | 3.92 | 3.92 | **3.85** |
| Effective approaches for strengthening post-surgical follow-up in the community | 26 | 3.77 | 3.81 | 3.77 | 4.00 | **3.84** |
| Effective mechanisms for client feedback and social accountability for respectful surgical MH/FP care | 27 | 3.56 | 3.93 | 4.00 | 3.85 | **3.83** |
| Best practices for defining and monitoring patient and family experiences of surgical MH/FP care | 27 | 3.65 | 3.85 | 3.67 | 3.89 | **3.77** |
| Impacts of COVID-19 on surgical MH/FP services' availability, affordability, and use | 27 | 3.85 | 3.67 | 3.15 | 3.37 | **3.51** |

